# Superspreaders and lockdown timing explain the power law dynamics of COVID-19

**DOI:** 10.1101/2020.07.23.20160531

**Authors:** Alexei Vazquez

## Abstract

Infectious disease outbreaks are expected to grow exponentially in time when left unchecked. Containment measures such as lockdown and social distancing can drastically alter the growth dynamics of the outbreak. This is the case for the 2019-2020 COVID-19 outbreak, which is characterized by a power law growth. Strikingly however, the power law exponent is different across countries. Here I illustrate the relationship between these two extreme scenarios, exponential and power law growth, based on the impact of superspreaders and lockdown strategies to contain the outbreak. The theory predicts a relationship between the power law exponent and the time interval between the first case and lockdown that is validated by the observed COVID-19 data across different countries.

---

Research from the late 90s and the 2000s uncovered the heterogeneity of social connectivity patterns, causing deviations from common expectations [1–4]. Superspreaders are the manifestation of this heterogeneity in the contest of infectious disease outbreaks: most infected individuals infect a few other people, but a few infected superspreaders infect many people [5]. More precisely, superspreaders are primary cases generating a number of secondary cases much larger than the median of secondary cases generated by all infected individuals. The existence of superspreaders was noticed in the 2002-2004 severe acute respiratory syndrome (SARS) outbreak, as well as in the 2012 Middle East respiratory syndrome (MERS) outbreak. It is then not surprising that superspreaders have been observed in the ongoing COVID-19 outbreak [6–8].

In an earlier work, I investigated the influence of superspreaders on infectious disease outbreaks [9]. These analyses showed that superspreaders can lead to a new type of infectious disease dynamics that is better described by a power law growth. Recent reports indicate that the COVID-19 outbreak is in fact better fitted by a power law growth rather than an exponential growth [10, 11]. Here I provide further support for the power law growth by linking the power law exponent with the timescales of the outbreak.

Spreading processes are generally modelled using differential equations [3], generating functions [12] or branching process formulations [9, 13]. I will adopt the branching process formulation because it provides an intuitive understanding of how the superspreaders and the lockdown affect the outbreak dynamics. Figure 1A shows a schematic representation of a causal tree of disease transmission associated with an epidemic outbreak. The root of the tree is patient zero. Every other node represents an individual that was infected during the outbreak. Each link represents the transmission of the disease from one individual (the primary case) to another individual (the secondary case).

**FIG. 1.**
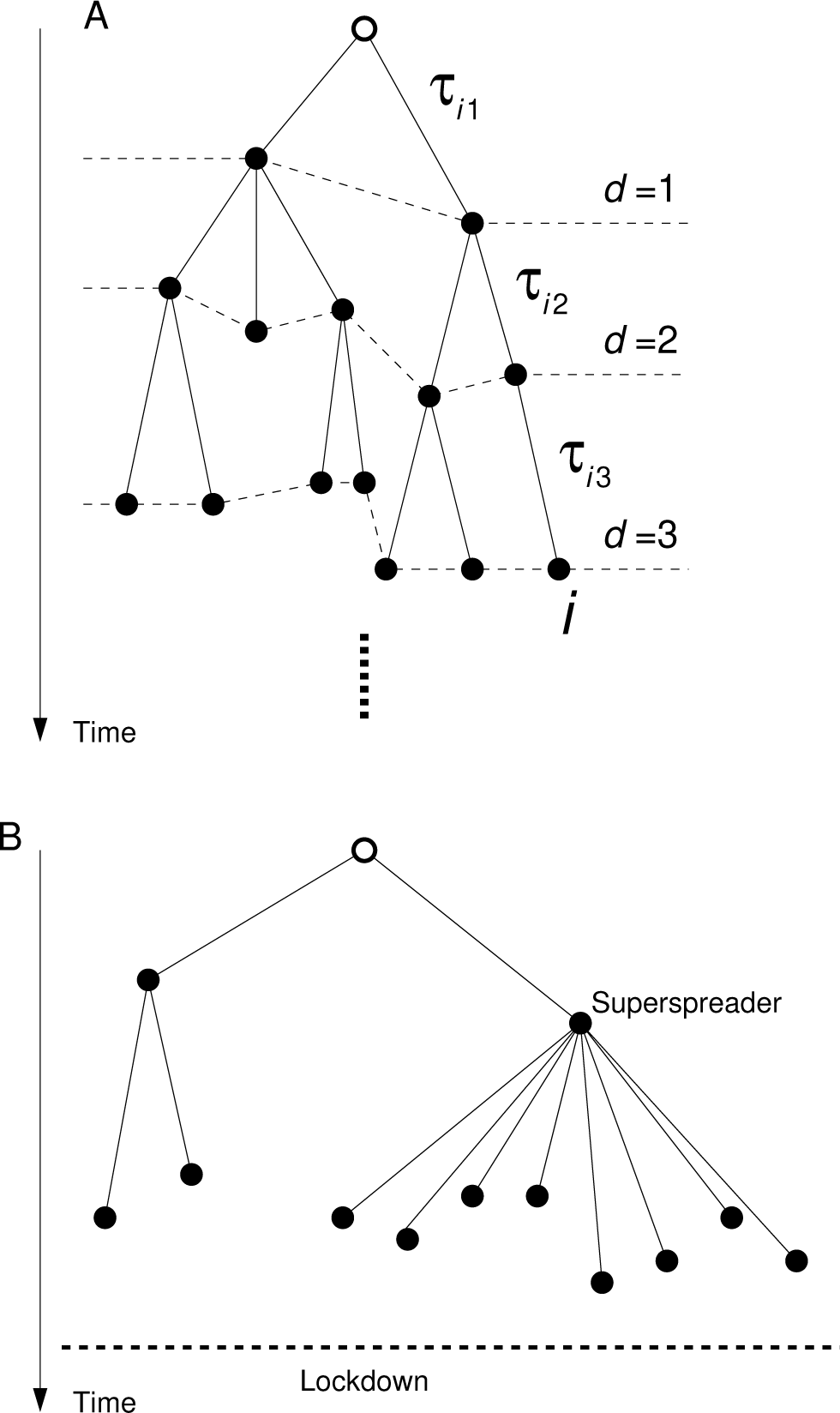
Causal tree of disease transmission of an infectious disease outbreak. A) Typical topology leading to an exponential growth. B) Typical topology leading to a power law growth. The comparison of the two panels highlights the distinctive difference between the trees topologies.

At the first generation we expect *R*_0_ infected individuals, where *R*_0_ is the average number of infectious generated by patient zero. At the second generation we expect *R*_0_*R* infected individuals, where *R* is the average number of infectious generated by patients other than patient zero. By applying the latter rule recursively we will expect

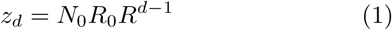

infected individuals at the generation *d* of the disease transmission tree. Here *N*_0_ represents the number of patients zero. *N*_0_ can be 1 or larger depending how many individuals got infected from the original source, *e*.*g*. an animal host or another country.

Notice that I have made an explicit distinction between patient zero and any other infected individual. This is done to take into consideration that the process of disease transmission introduces a bias in the connectivity statistics of infected individuals. For example, consider a model where each individual *i* is in close proximity with other individuals at a rate *λ*_*i*_, measured in proximity contacts per unit of time. Patient zero is an individual selected with uniform probability 1*/N*, where *N* is the population size. Each time patient zero is in close proximity with another individual, the latter is infected with probability *r*, where *r* is the probability of disease transmission upon contact. If *T* = 1*/γ* is the infectious period, the inverse of the recovery rate *γ*, and *β*_*i*_ = *rλ*_*i*_ is the rate of disease transmission, then patient zero will generate the average number of secondary cases given by

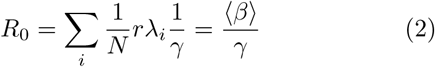

In contrast, subsequent infected individuals are not selected at random from the population. In a fully mixed population, when an infected case is in close proximity to another individual, the probability that it is the *i*-th individual is *λ*_*i*_*/N* ⟨*λ*⟩. In turn, if *i* gets infected it will transmit the disease at a rate *rλ*_*i*_ = *β*_*i*_. Therefore

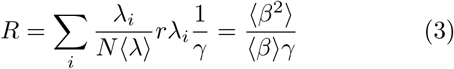

The latter result is similar to what obtained in static networks [3, 9, 12], where the reproductive number is proportional to the ratio between the first and second moments of the degree distribution. The mapping between the contact dynamics and static network representation can be done after taking into account that the number of contacts in a time interval *T* follows a Poisson distribution, resulting in the degree distribution

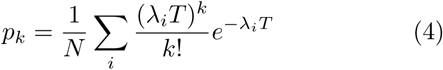

From the latter equation we can calculate the first and second moment of the degree distribution, obtaining ⟨*k*⟩ = ⟨*λ*⟩ *T* and ⟨*k*(*k* − 1) ⟩ = ⟨*λ*^2^⟩ *T/* ⟨*λ*⟩, respectively. Substituting these values into equations (2) and (3) we finally obtain

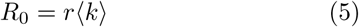

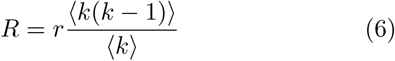

which is the average reproductive numbers of the root and other nodes as obtained in the static network representation [3, 9, 12]. The −1 in equation (6) excludes the link from where the node gets infected.

To determine the number of new infectious at a given time, we need to make a mapping from generation to infection time. For an infected individual at generation *d*_*i*_, the infection time equals the sum of *d* transmission times.

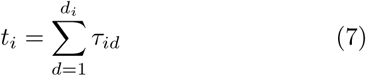

where *τ*_*id*_ is the disease transmission time from the primary case at layer *d* − 1 to the secondary case at layer *d*, within the path from patient zero to node *i* (see Fig. 1A). If *g*(*t*) is the probability density function of the disease transmission time from a primary to a secondary case, then the probability distribution that an infected individual at generation *d* gets infected at time *t* is given by the *d* − 1 order convolution of *g*(*t*), denoted by

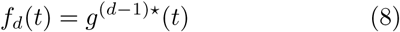

If infected individuals are removed at a rate *γ*, then the generation time distribution is exponential, *g*(*t*) = *γe*^−*γt*^ with average generation time *T* = 1*/γ*. In this particular case we can derive an analytic expression

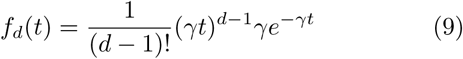

Now we are ready to complete the mapping from generations to time. From equations (1) and (9) it follows that the average number of new infections at time *t* is given by

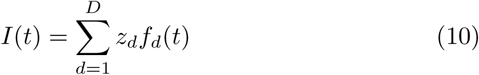

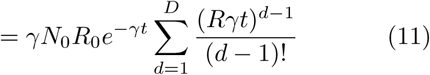

where *D* is the final generation, when the outbreak ends due to natural extinction or interventions strategies. A formal derivation of equation (11) can be found in [9, 13].

To understand the impact of superspreaders and lockdown, let us have a look at the two trees of disease transmission in Fig. 1A and B. In Fig. 1A most individuals transmit the disease approximately to the same number of other individuals, and the chain of transmissions extends in this manner over several generations. When *D* is very large, equation (11) represents the Taylor expansion of the exponential, resulting in

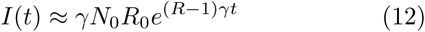

This approximation is valid for *γtR* ≪ *D*. If *R >* 1 then the outbreak grows exponentially over time, else if *R <* 1 the outbreak decays exponentially. This is the canonical expectation of infectious disease dynamics. In this scenario the key quantity is the reproductive number *R* and interventions strategies are focused on bringing it below 1.

However, there are a number of assumptions behind equation (12) that make it inadequate to model all infectious diseases outbreaks. First, we have just assumed that *D* is large, *i*.*e*. that the free spreading of the disease goes over several generations. That would typically be true for infectious diseases with mild symptoms such as the common cold, but it is not the case for COVID-19. The mortality and hospitalization rate of COVID-19 infections have led governments to impose strict lockdown measures. As a consequence, the tree of disease transmission is truncated after a few generations, as shown in Fig. 1B.

Second, there are superspreaders. The reproductive number of individuals other than patient zero is proportional to the ratio between the second and first moments of the distributions of contact rates. If there is a wide variation of contact rates in the population, ⟨*β*^2^⟩ ≫ ⟨*β*⟩, and the probability of disease transmission upon contact is high, then according to equation (3) *R* will be large. In the specific case of the COVID-19 outbreak, the probability distribution *q*_*k*_ of secondary cases *k* generated by a primary case has a fat tail. The fat tail is well approximated by the power law 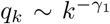, where *γ*_1_ ≈2 based on world-wide data [14]. The exponent *γ*_1_ = 2 is the expectation for a contagion spreading in networks with multiple topologies and therefore it has a theoretical foundation [15]. When *γ*_1_ ≤ 2, the value of *R* = Σ_*k*_ *q*_*k*_*k* is ill-defined, it will increase as more disease transmissions are allowed. When *γ*_1_ ⪆ 2 the value of *R* is well defined but it will be very large, diverging as *γ*_1_ → 2.

When these two elements are taken into consideration, a small number of generations *D* and the existence of superspreaders, then equation (11) is better approximated by [9]

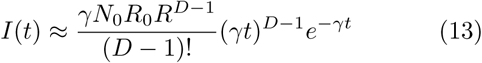

This approximation is valid for *γtR* ≫ *D*. In short, the number of infected cases from one generation to the next can increase so dramatically that the number of new daily infectious will be dominated by the time the individuals at the last generation get infected. The power law behaviour predicted by equation (13) is so different from the readily explainable exponential behaviour (12) that it has been neglected for 14 years.

Equation (13) makes a testable prediction, that the exponent of the power law growth, *I*(*t*) ∼ *t*^*α*^, depends on *D*,

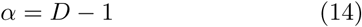

In turn, *D* can be estimated as the number generations of disease transmission from patient zero to the implementation of lockdown,

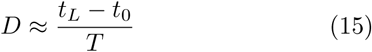

where *t*_*L*_ is the time when the lockdown was implemented and *t*_0_ is the time when the first case was reported. *T*, as before, is the average time from being infected to disease transmission. Combining equations (14) and (15) then yields

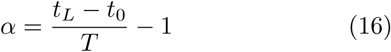

To test equation (16), I collected data for the observed power law exponents. Singer has conducted an extensive analysis of the fitting of a power law or logistic growth to the plot of new infections as a function of time [11]. Here, I focus on those countries were the power law fit was dimmed a better fit than a logistic function. As an example, Fig. 2 shows the data for the Netherlands. The log-log plot emphasizes the power low growth of the outbreak before lockdown (Fig. 2, red line). The power law exponents are reported in the Table 3 of Ref. [11].

**FIG. 2.**
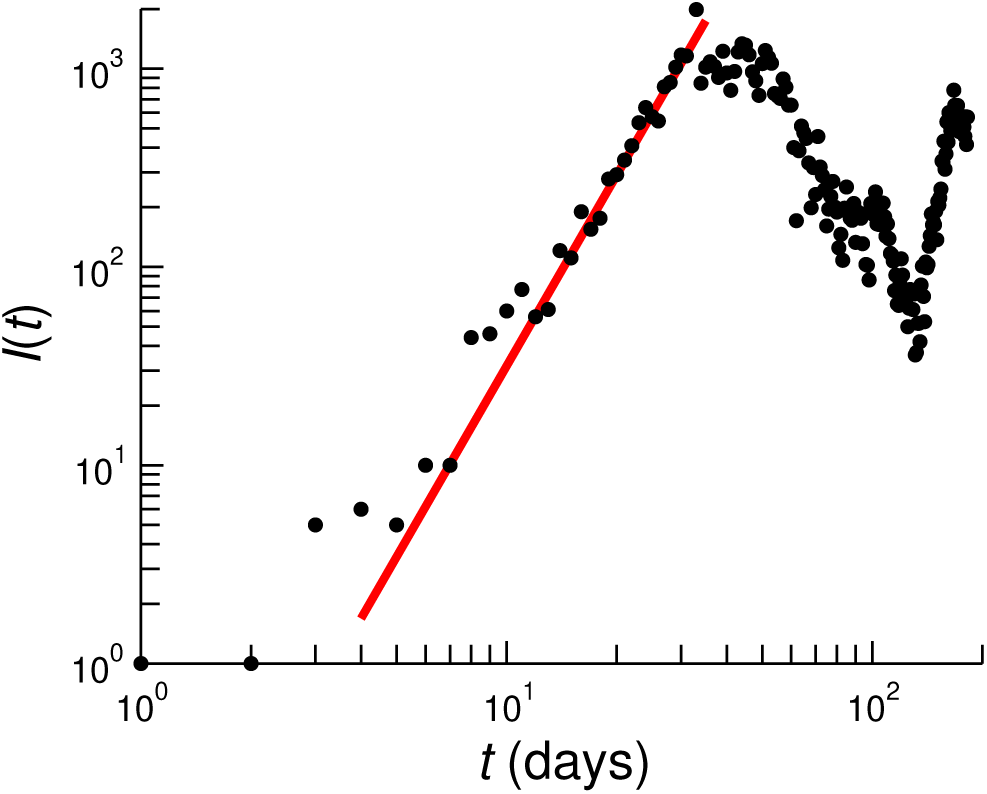
Number of new infections in the Netherlands as a function of time, measured in days from February 28th, when first case was reported. Based on data retrieved from the World Health Organization (WHO) website. The red line represents a power law growth ∼ *t*^3.2^.

In parallel, I have estimated the power law exponent using equation (16). To this end, the time of first confirmed case were retrieved from the World Health Organization (WHO) website at https://covid19.who.int. Except for China, that was assumed as 8th of December, when the first suspected case in Wuhan was reported having symptoms of coronavirus. The time of lockdown was assumed to be the 20th of March for all countries but China, as reported in [16] and available at https://www.politico.eu/article/ europes-coronavirus-lockdown-measures-compared. For China we assumed the 1st of January, when the Wuhan market was closed. *T* was estimated as the COVID-19 incubation time, which is approximately 5 days [17]. Based on these parameter estimates we obtain the exponent values predicted by equation (16), which are in very good agreement with the exponents obtained by Singer from a direct fit to the *n*(*t*) vs *t* data (Fig. 3).

**FIG. 3.**
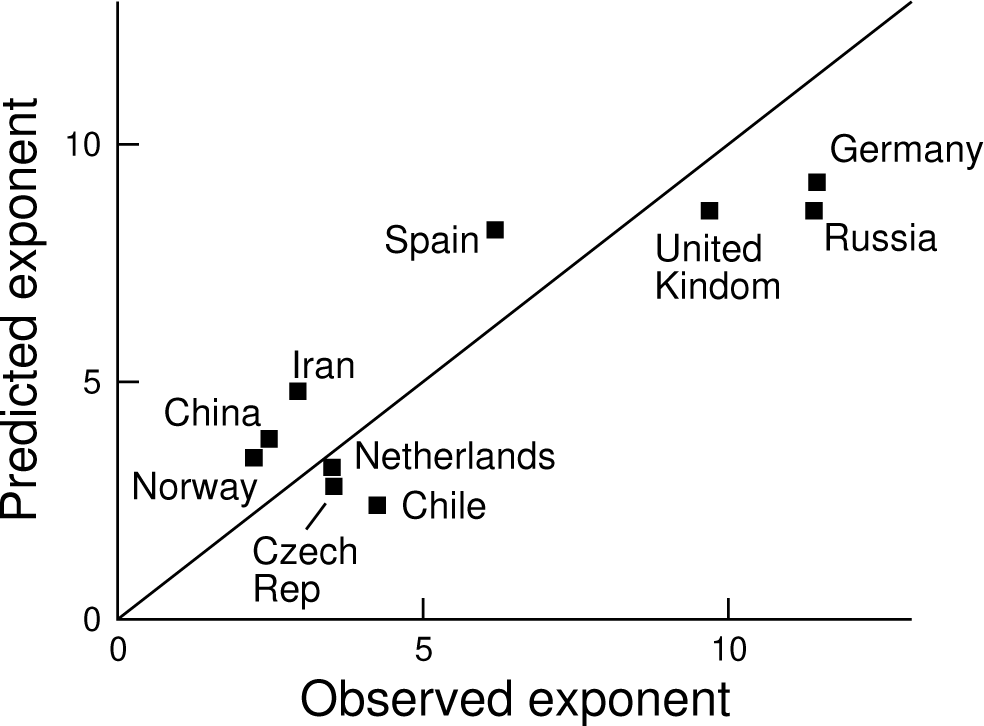
Relationship between the observed and expected power law exponent. The points are based on observational data for the COVID-19 outbreak in the indicated countries. The line represents the prediction being equal to the observation.

In conclusion, the power law dynamics of the COVID-19 outbreak is a validation of the new power law of infectious disease spreading [9]. This is further demonstrated by the relationship between the power law exponent and the time interval between first case and lockdown. Furthermore, it can be shown that the power law growth persists even if there are degree correlations [18] or multiple types of spreaders [19]. Again this study also underscores the crucial importance of early lockdown timing in the control of an infectious disease in a population with superspreaders.

These results are relevant for the management of coronovirus outbreaks or any other outbreak with superspreaders. First, the common assumption of exponential growth/decay needs to be revised. Specially when it comes to estimate the basic reproductive number from the plot of new infections as a function of time. Second, the theory explains why the power law exponent is variable across countries. The value of the power law exponent contains information about the number of generations the outbreak went through.

## Data Availability

All data is reported in the manuscript.

## ACKNOWLEDGEMENTS

This work was supported by Cancer Research UK C596/A21140.

